# A clinical decision rule to reduce unnecessary radiographs in pediatric finger injuries

**DOI:** 10.1101/2025.05.23.25327759

**Authors:** Sara Suarez-Cabezas, Elisa M. Molanes-Lopez, Almudena García García-Galan, Eva Sanavia Moran, Jose Meizoso Lopez, Begoña Perez-Moneo

**Affiliations:** Hospital Universitario Infanta Leonor, Servicio de Pediatría. Madrid, Community of Madrid, Spain; Universidad Complutense de Madrid, Departamento de Estadística e Investigación Operativa, Facultad de Medicina, Madrid, Community of Madrid, Spain; Universidad Complutense de Madrid, Departamento de salud pública y materno-infantil. Facultad de Medicina. Madrid, Community of Madrid, Spain

## Abstract

**Objective:** To develop and validate a Clinical Decision Rule (CDR) for paediatric finger trauma that reduces unnecessary radiographs while ensuring accurate identification of clinically significant fractures.

**Design:** Observational, retrospective, single-center study. Medical records of patients aged 0–16 years presenting with finger trauma were reviewed to develop a CDR. Multivariate logistic regression analysis was performed to identify predictive variables. A diagnostic test study was then carried out to assess the performance of the developed CDR.

**Setting:** paediatric emergency department of a secondary-level hospital.

**Participants:** A total of 462 injured fingers from 411 patients.

**Main Outcome Measures:** Sensitivity, specificity, positive and negative predictive values (PPV and NPV) for significant fractures; Absolute Risk Reduction (ARR), Relative Risk Reduction (RRR), and the Number Needed to Treat (NNT) for the reduction in X-rays requested and their 95% confidence interval (95% CI).

**Results:** A total of 411 patients (462 injured fingers) were included, with a mean age of 10.13 years (standard desviation 3.70) and 57.9% being male. Overall, 8.01% of cases had a fracture, with a significantly higher prevalence in males (10.38% vs. 4.95%, p = 0.0328). Multivariate logistic regression identified deformity (adjusted Odds Ratio (OR): 2.33×10^2^), mid/distal phalanx injury (OR: 15.40), impaired function (OR: 9.48), and hyperextension/flexion mechanism (OR: 5.37) as significant predictors. A CDR was developed with 100% sensitivity (95% CI 65.55 to 100) and 100% NPV (95% CI 98.11 to 100) for detecting clinically significant fractures. Applying the rule reduce 41.87% (ARR 95% CI: 36.20 to 47.55%) of radiographs, with an estimated annual cost saving of €2,650.

**Conclusions:** This CDR is a reliable tool for identifying patients at risk of significant fractures in finger trauma. It safely reduces radiograph use, decreases radiation exposure, and optimizes healthcare costs. Further external validation through prospective studies is needed.

**KEY MESSAGES:** *What is already known on this topic:* No validated clinical decision rule exists for paediatric finger trauma, despite the high frequency of such injuries and the frequent overuse of radiographs. While clinical decision rules in other anatomical regions (e.g., ankle, knee) have proven effective in reducing unnecessary imaging, similar tools have not been validated for hand trauma in children.

*What this study adds:* This study proposes a clinical decision rule for paediatric finger trauma, based on a retrospective analysis. The rule demonstrated high diagnostic performance, with 100% sensitivity and negative predictive value for significant fractures, and could potentially reduce radiograph use by over 40%.

*How this study might affect research, practice or policy:* Although external and prospective validation is still required, this clinical decision rule shows promise as a future tool to optimise decision-making in paediatric finger trauma. If validated, it may contribute to reduce unnecessary radiographs, improving emergency department workflow, and minimising radiation exposure in children.

## INTRODUCTION

Hand injuries are among the most common traumatic events in the pediatric population(1–3). Together with wrist and hand injuries, they account for approximately 1.5% of emergency department consultations(2). In our center, these injuries represent 17% of the extremity trauma cases treated in the Pediatric Emergency Services (PES), making them the second most frequent location after ankle injuries(3).

Most of these injuries are contusions and sprains, with fracture incidence ranging from 21% to 65%(4–6). However, a significant proportion of these fractures are considered non-displaced and can be effectively managed with immobilization through finger splinting, leading to full recovery without sequelae(6–9).

To optimize the use of radiographs, various clinical decision rules (CDRs) have been developed. Notable examples include the Ottawa criteria for ankle injuries and the Low-Risk Ankle Rule, which have been shown to reduce the number of radiographs ordered by up to 60%(10–12). Similarly, the Ottawa criteria for knee injuries have demonstrated similar effectiveness(13). However, despite the high frequency of hand finger trauma in PES, no specific CDR has been validated for this location to reduce unnecessary radiograph use.

This study aims to develop a CDR for detecting significant fractures following hand finger trauma, with the goal of decreasing the number of radiographs ordered and optimizing clinical management for these patients.

## MATERIALS AND METHODS

A retrospective, observational, single-center study was conducted at a secondary-level hospital, following approval from the Ethics Committee for Research at our institution.

### Study Population

The electronic medical records of all patients aged 0 to 16 years who presented to the PES for trauma of any of the hand fingers were reviewed. The exclusion criteria were: absence of radiograph, penetrating trauma, complete or partial finger amputation, simple erosion or wound without significant trauma, pre-existing or concurrent conditions that increased the risk of fractures, and previous pathology of the affected finger.

### Variables Collected

Age, sex, number of the injured finger, mechanism of injury, laterality, injury location, presence of edema, bruising, functional impairment, deformity, instability and/or skin injury, presence of fracture on the radiograph, and diagnosis of significant fracture.

### Definition of Significant Fracture

Following a review of the literatura(7–9,14), significant fractures were defined as intra-articular fractures (except for avulsion fractures of the volar plate affecting less than one-third of the articular surface), fractures with deformity (angular, axial, or rotational), displaced fractures, and joint dislocations or subluxations.

### Statistical Analysis

Categorical variables were described using percentages, while continuous variables were expressed as means and standard deviations (SD) if they followed a normal distribution (assessed using the Kolmogorov-Smirnov and/or Shapiro-Wilk tests), or as medians and interquartile ranges (IQR) otherwise.

A binary logistic regression analysis was performed to identify predictive variables for significant fractures. All the independent variables that could influence the presence of a significant fracture were selected, including: sex, age, mechanism of injury, affected finger number, injury location, and physical examination findings. The dependent variable was the diagnosis of significant fracture based on the radiographic result (anteroposterior and lateral x-ray).

Several predictive models were constructed, calculating sensitivity (S), specificity (E), positive predictive value (PPV), and negative predictive value (NPV) with a 95% confidence interval (95% CI) for each, using the radiographic result as the gold standard. A ROC curve analysis was conducted to select the model that achieved 100% sensitivity for detecting significant fractures. Additionally, an estimate was made of the reduction in the number of radiographs requested by applying the proposed CDR, calculating the Absolute Risk Reduction (ARR), Relative Risk Reduction (RRR), and the Number Needed to Treat (NNT), along with their 95% CIs.

### Economic Analysis

An economic analysis was performed based on the estimated reduction in the number of hand finger radiographs and their cost at our institution, which was €9.47 per radiograph.

### Sample Size

The calculated sample size was 407 patients, aiming to detect risk factors associated with an Odds Ratio (OR) of 4 in a multivariate logistic regression adjustment, with the response variable being significant fracture with a relative frequency of 4%(3). An alpha error of 0.05 and a power of 80% were assumed. The risk factor was considered binary with a balanced design (risk proportion pi=0.5), such that the square of the multiple correlation coefficient with the other risk factors was R^2^ = 0.40(15). To achieve this sample size, a retrospective review of electronic medical records from April 2019 to March 2020 was planned.

## RESULTS

A total of 462 fingers affected by trauma were included, corresponding to 411 patients (Figure 1). The mean age was 10.13 (SD = 3.70), with a predominance of males (57.91%) compared to females in the reference population, yielding a male-to-female prevalence ratio (PR) of 1.30 (95% CI 1.07 to 1.58, p 0.0081).

**Figure 1.**
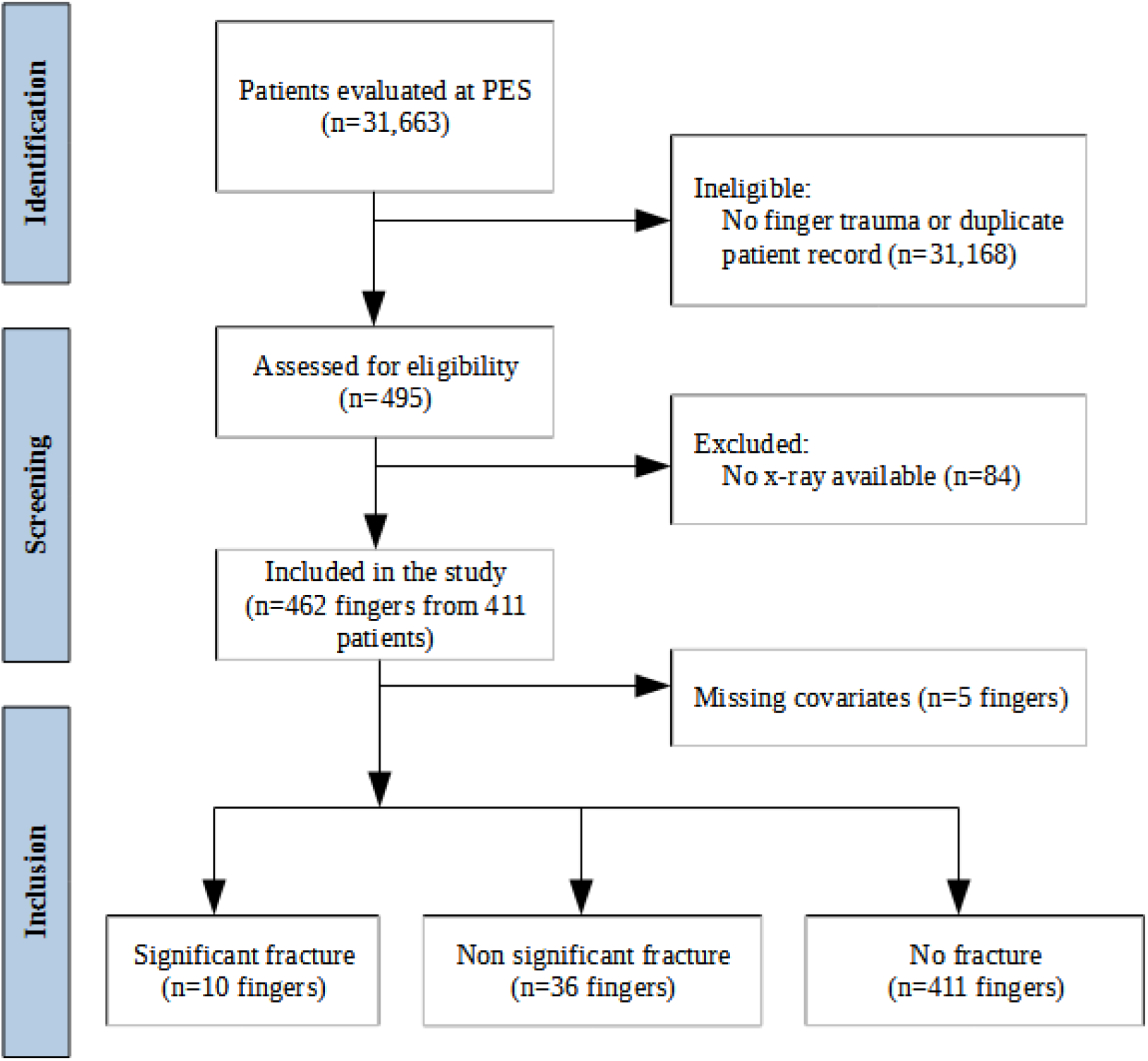
TRIPOD flow diagram: patient identification and selection.

The remaining baseline characteristics are described in Table 1.

**Table 1.**
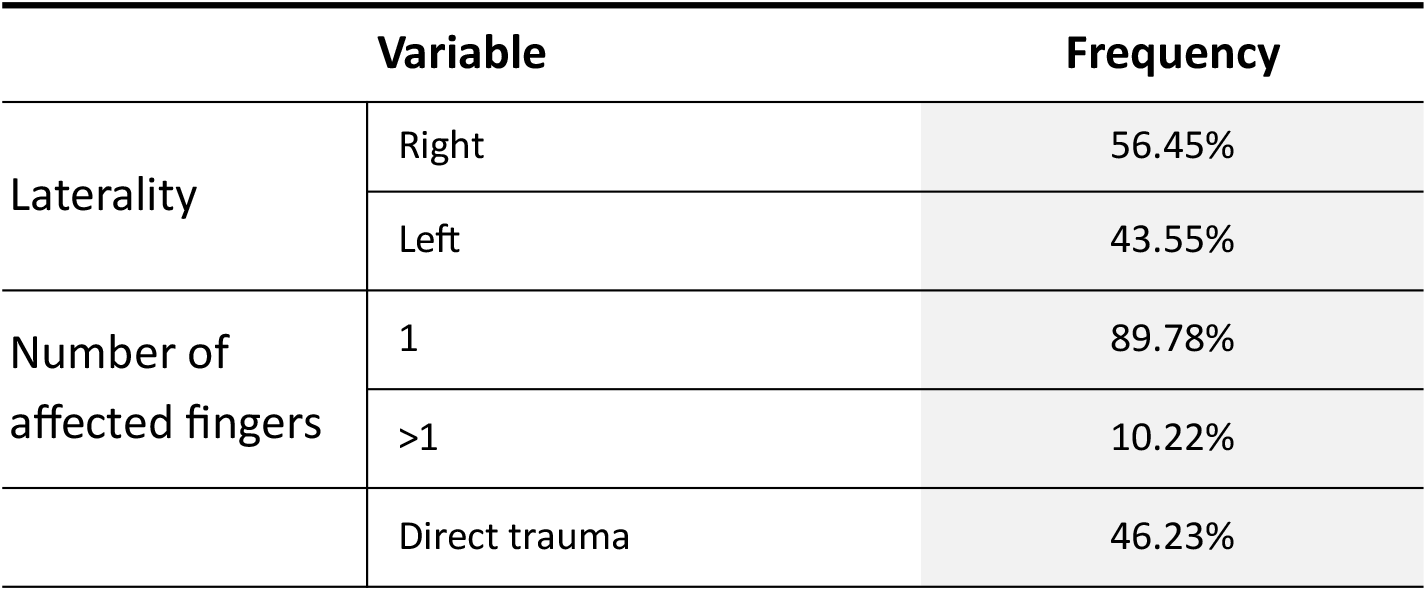
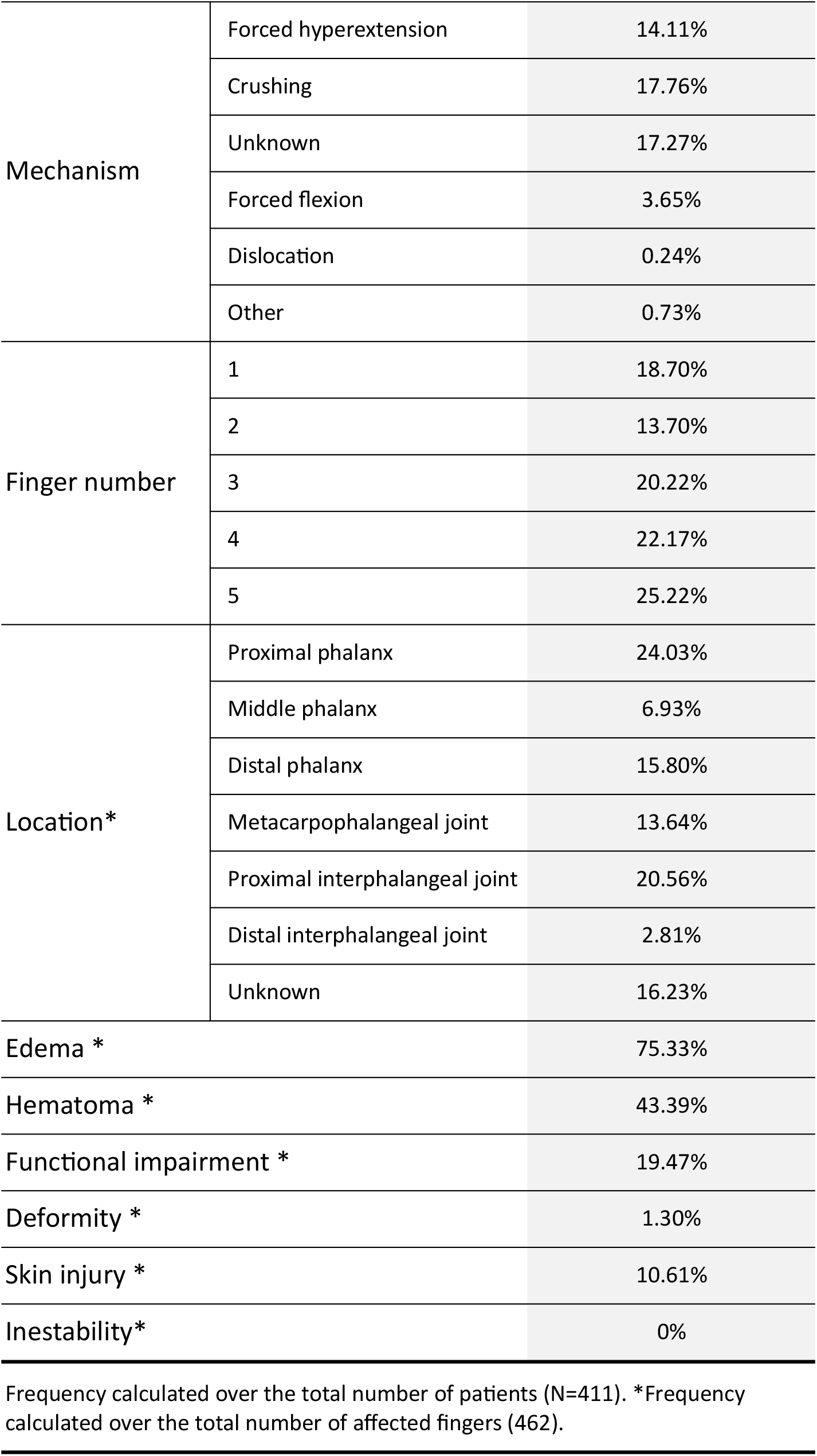
Baseline characteristics of the patients included in the study.

A fracture was detected in 8.01% of cases. The prevalence was significantly higher in males (10.38%) than in females (4.95%), with a male-to-female prevalence ratio (PR) compared to the reference population of 2.10 (95% CI: 1.04 to 4.23, p 0.0328). The prevalence of significant fractures was 2.16%, with the gender difference still evident (3.46% in males vs. 0.50% in females), yielding a male-to-female PR of 6.99 (95% CI: 0.89 to 54.74, p 0.0481).

Furthermore, the incidence of fractures was higher in the 5th finger, followed by the 3rd and 4th fingers, as shown in **Table 2**.

**Table 2.** Fracture frequency by finger number, calculated from a total of 37 fractures.

| Number of finger | Fracture frequency |
| --- | --- |
| 1 | 10.81% |
| 2 | 2.70% |
| 3 | 27.03% |
| 4 | 18.92% |
| 5 | 40.54% |

### Development of the Clinical Decision Rule

A univariate and multivariate logistic regression model was fitted, including only male patients, as the inclusion of females worsened model performance. The dependent variable was the presence of fracture, and the independent variables were: age, mechanism of injury, affected finger, injury location, presence of edema, hematoma, functional impairment, skin injury, and deformity. Instability was excluded from the analysis, as no patient presented this finding on physical examination.

The final multivariate model, obtained using any of the stepwise methods (backward, forward, or bidirectional), identified the presence of deformity as the factor most strongly associated with fractures, with an adjusted OR of 2.33×10^9^ (Table 3). No significant association was found between age, edema, hematoma, or skin injury and the presence of fracture. Non-binary variables, such as mechanism of injury, affected finger number, and injury location, were transformed into binary variables prior to inclusion in the model, based on previously obtained descriptive data.

**Table 3.** Results of univariate and multivariate binary logistic regression models.

| Variable | Crude OR<br>from the<br>univariate<br>models<br>(p-value) | Adjusted OR<br>from the full<br>model with<br>n=246<br>(p-value) | Adjusted OR<br>from the final<br>model with<br>n=253<br>(p-value) |
| --- | --- | --- | --- |
| Deformity (n=260) | $1.59 \times 10^8$<br>($<0.0001$) | $2.10 \times 10^9$<br>($<0.0001$) | $2.33 \times 10^9$<br>($<0.0001$) |
| Injury to middle or distal phalanx (n=260) | 2.75<br>(0.0196) | 17.92<br>($<0.0001$) | 15.40<br>($<0.0001$) |
| Functional impairment (n=253) | 4.61<br>(0.0005) | 6.98<br>(0.0003) | 9.48<br>($<0.0001$) |
| Hyperextension or hyperflexion mechanism (n=260) | 1.85<br>(0.1931) | 4.73<br>(0.0137) | 5.37<br>(0.0066) |
| Injury to index finger (n=260) | $6.44 \times 10^{-8}$<br>($<0.0001$) | $1.65 \times 10^{-8}$<br>($<0.0001$) | $1.70 \times 10^{-8}$<br>($<0.0001$) |
| Age (n=260) | 1.04<br>(0.4991) | 1.06<br>(0.5058) | N/A |
| Edema (n=255) | 8.47<br>(0.0383) | 3.44<br>(0.3009) | N/A |
| Hematoma (n=253) | 1.77<br>(0.1874) | 1.07<br>(0.9046) | N/A |
| Skin injury (n=260) | 0.73<br>(0.6821) | 0.96<br>(0.9671) | N/A |

Based on the variables identified as significant in the final model, the CDR described in Figure 2 was developed, achieving a sensitivity (S) of 100% for the detection of significant fractures.

**Figure 2.**
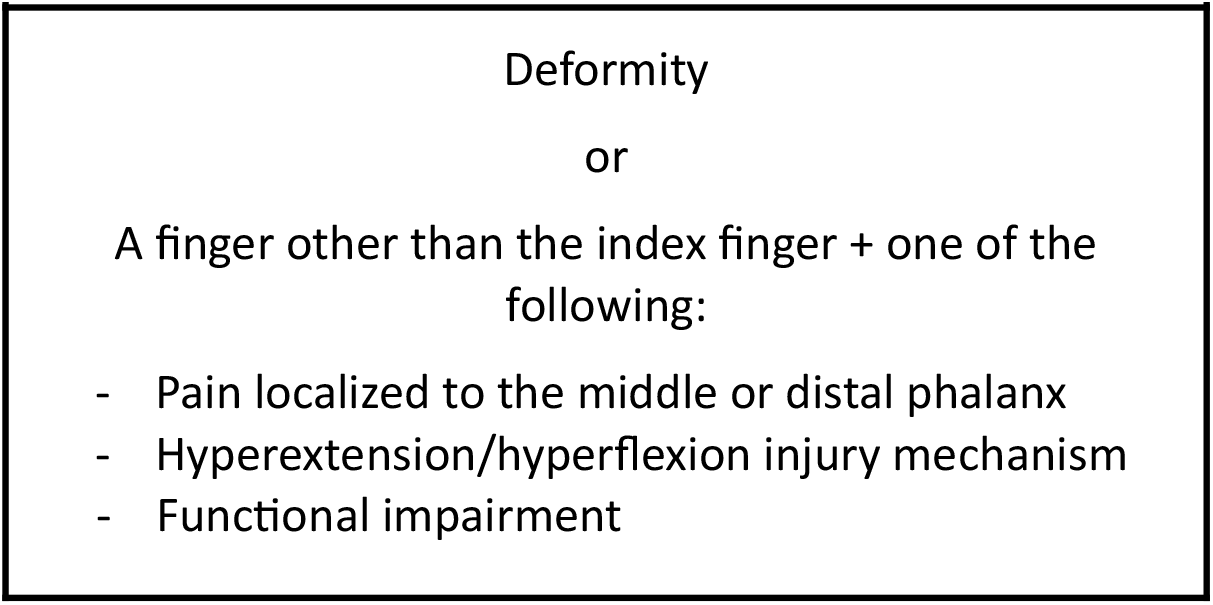
Clinical Decision Rule

The ROC curve associated with the final model is shown in Figure 3.

**Figure 3.**
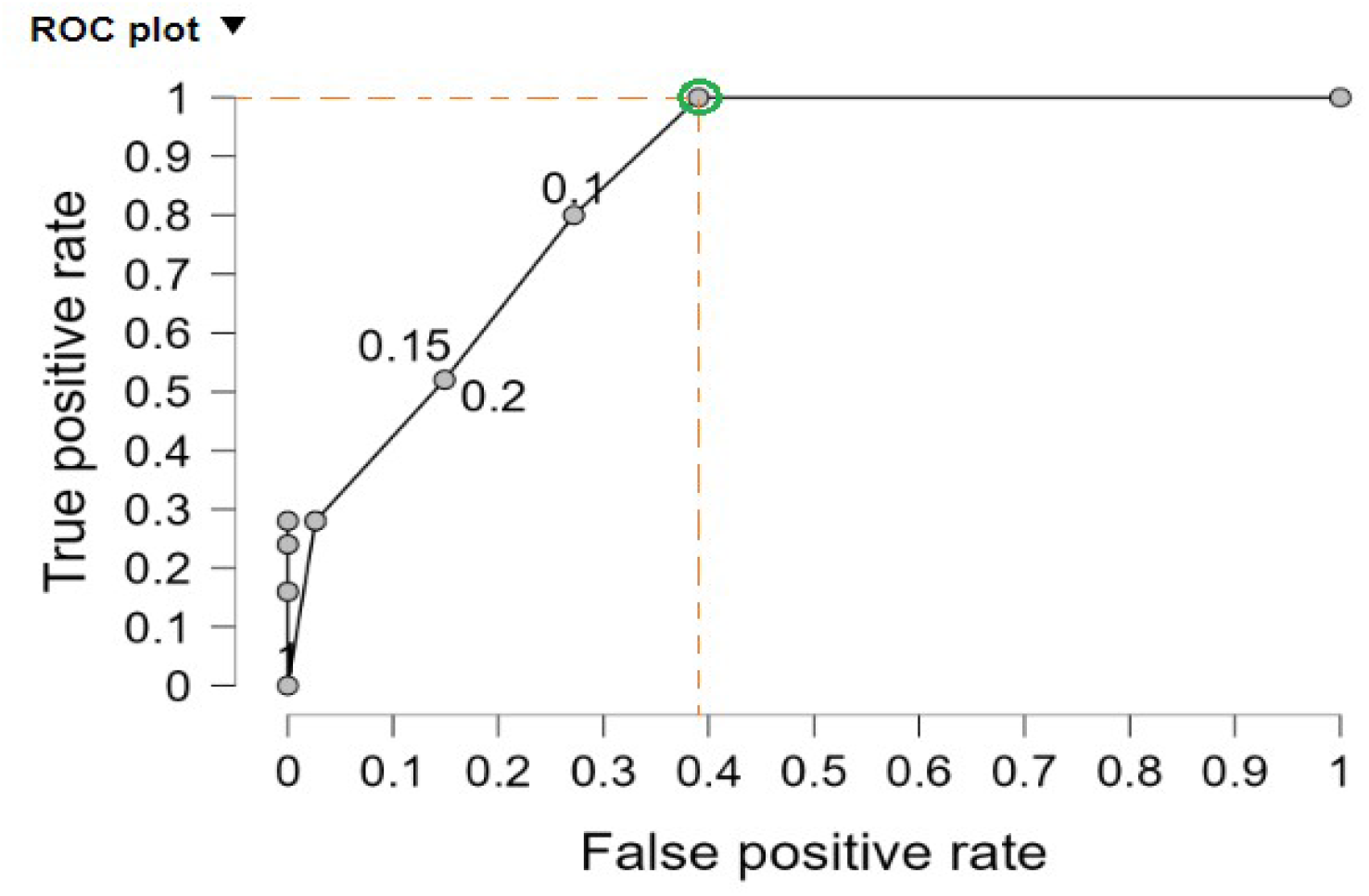
ROC curve of the selected model used to develop the CDR.

### Clinical Decision Rule Analysis

When comparing the CDR to radiographic findings (gold standard), and including both male and female patients, the rule demonstrated a sensitivity of 100% (95% CI: 65.55 to 100), a specificity of 55.70% (95% CI: 50.96 to 60.35), a PPV of 4.81% (95% CI: 2.46 to 8.92), and a NPV of 100% (95% CI: 98.11 to 100) for the detection of significant fractures.

Detailed results by sex for both overall and significant fractures are shown in Table 4.

**Table 4.**
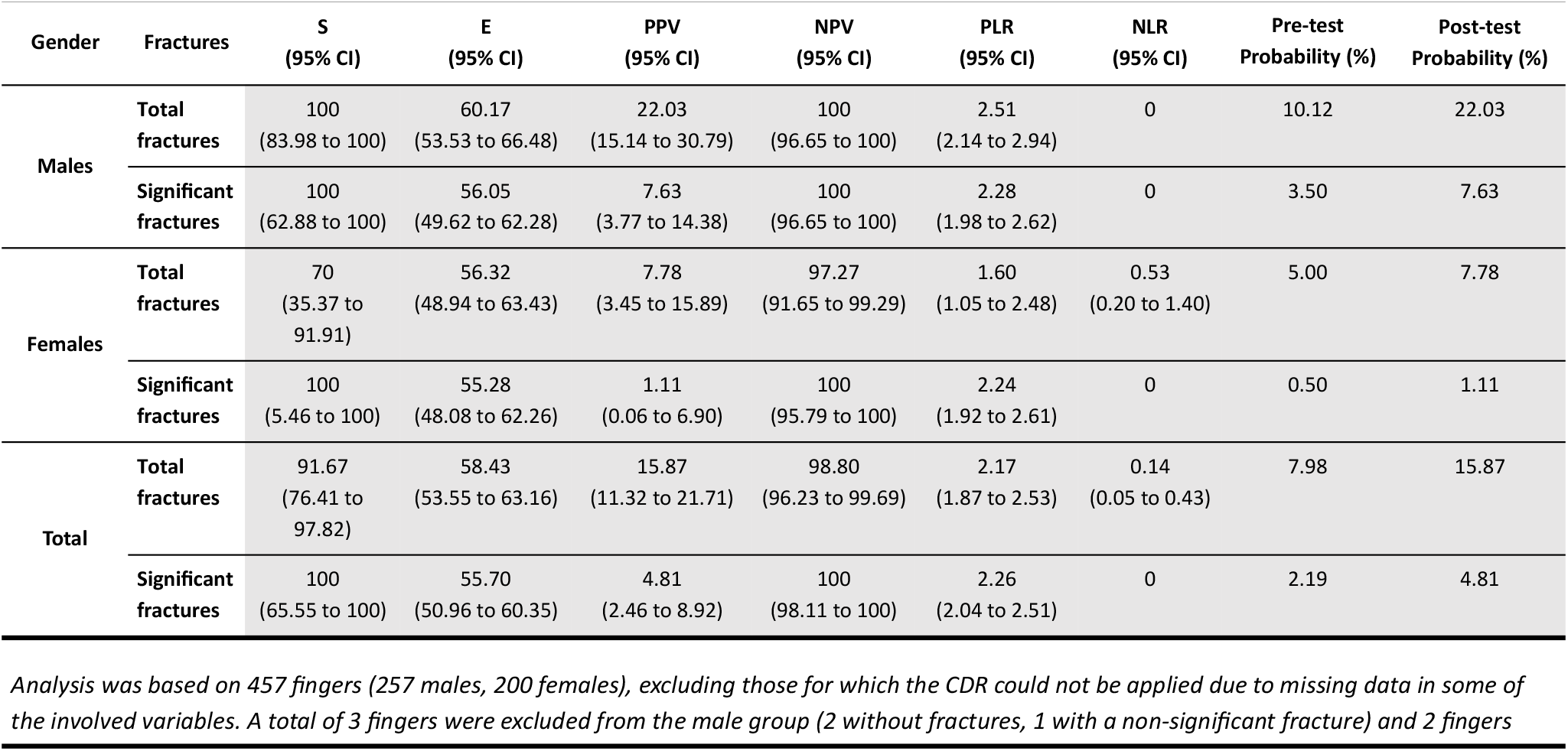
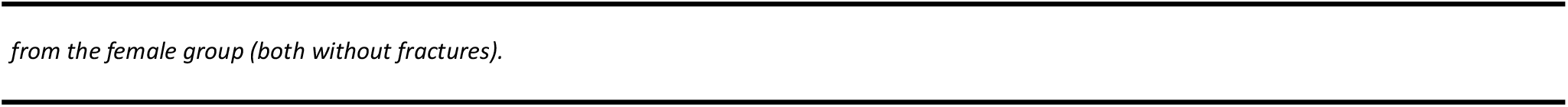
Diagnostic accuracy of the CDR for detecting total or significant fractures stratified by gender.

### Reduction in the number of X-rays

Considering that, in usual practice, an X-ray is requested in 90% of patients with hand finger trauma(3), and that in our sample, 48.03% of patients met the CDR, the ARR was estimated at 41.87% (95% CI: 36.20 to 47.55%), RRR at 46.58% (95% CI: 40.58 to 51.96%), and NNT at 2.39 (95% CI: 2 to 3).

In practical terms, considering that approximately 700 patients are treated annually in our center for finger trauma, the application of this CDR could reduce the number of X-rays requested by 41.87%, equivalent to 293 fewer X-rays per year. This would result in an annual economic saving of approximately €2,650.

## DISCUSSION

This study develops a CDR for hand finger trauma that safely reduces the number of X-rays requested by 40%, with a sensitivity of 100% and a negative predictive value of 100% for the detection of significant fractures.

The fracture rate in our study was 8%, similar to what has been reported in previous reviews(3), but lower than the 21-65% range described in other studies(4–6,14). This discrepancy could be explained by an increase in consultations for minor injuries in the PES, a trend already observed by other authors(16,17).

Furthermore, we confirm a higher frequency of trauma in men (58%), consistent with previous studies that report rates of 56-61%(5,18). This male predominance is accentuated when considering total fractures and, especially, significant fractures, a factor that had not been considered in other studies.

Regarding the location of the injuries, we found that the 5th finger was the most affected (40%), followed by the 4th and 3rd fingers. This pattern, not previously described, may be related to reduced force in the fingers further from the grip. While Vadivelu et al.(4) also reported a higher frequency of fractures in the 5th finger (52%), the frequency in the 1st finger was lower in our study (10%). It is worth noting that the fracture rate found in the 2nd finger was almost anecdotal, making it a protective factor in our CDR.

### Development and Validation of the Clinical Decision Rule

To date, no validated CDR exists to reduce the number of radiographs in cases of finger trauma, unlike other locations such as the ankle(10–12) or knee(13). Jawad et al.(19) highlighted the need for criteria that would minimize unnecessary radiographs, reporting a ratio of 1:23 between radiographs with clinical impact and those without.

Recently, Steiger et al.(14) proposed a CDR for significant fractures in the proximal interphalangeal joint following a hyperextension mechanism due to a ball-related trauma, achieving 100% S and 100% NPV. However, their approach was based on subjectively selected variables, potentially overlooking relevant predictive factors, and has not been validated. In contrast, our study develop a CDR applicable to any location and mechanism of injury, confirming that deformity is the strongest predictor of significant fractures.

### Clinical and Economic Implications

Our CDR demonstrated 100% S and 100% NPV for significant fractures, and 100% sensitivity and 100% NPV for total fractures in males, although not in females. This suggests that its application in females, where the proportion of significant fractures is so low, requires caution.

Implementing this CDR could reduce radiographs by 40%, comparable to results from criteria applied in other locations(10–12). This would not only decrease radiation exposure but also reduce waiting times in emergency departments and generate an estimated annual cost saving of €2,650 at our center.

### Limitations

Our study has several limitations. First, its retrospective design introduces potential biases in data collection and excludes patients with incomplete information. Additionally, the CDR has not been externally validated, so prospective studies are needed to confirm its efficacy.

## CONCLUSIONS

This study developed a clinical decision rule for finger trauma, which safely identifies patients at risk of significant fractures. This CDR, with a 100% negative predictive value, reduces the number of radiographs requested by 40%, thereby decreasing patient radiation exposure, shortening waiting times, and optimizing healthcare expenditure.

## Data Availability

All data produced in the present study are available upon reasonable request to the authors

